# Impact of 3D Printed Models on Shared Decision Making: A Cluster Randomized Controlled Trial

**DOI:** 10.1101/2025.01.27.25321192

**Authors:** Aimal Khan, Georgina E. Sellyn, Danish Ali, Zorays Moazzam, Hillary Samaras, Shannon McChesney, M. Benjamin Hopkins, Molly M. Ford, Roberta Muldoon, Timothy M. Geiger, Dann Martin, Daniel L. Chu, Kyle K. VanKoevering, Alexander T. Hawkins

## Abstract

**Importance:** Surgical patients report a lack of involvement in healthcare decisions and increased anxiety. 3D models serve as educational tools to encourage patient engagement, reduce anxiety levels, and aid understanding. We hypothesized that patients who receive pre-operative education with by 3D-printed anatomic models would perceive a higher involvement in decision making and experience lower anxiety levels versus “standard care.”

**Objective:** To determine the impact of 3D-printed models on shared decision making and patient anxiety during the pre-operative surgical consultation.

**Design:** Single-center cluster randomized control trial.

**Setting:** Colorectal surgery clinic at a tertiary hospital.

**Participants:** Adult (18 years or older) patients scheduled for partial or complete colon and/or rectal resection.

**Intervention/s:** Surgeons were cluster-randomized to counsel patients using a modular 3D-printed model or providing standard care during pre-operative clinic visits.

**Main outcomes:** The primary outcome was the patient’s perception of involvement in decision making, assessed using the validated Shared Decision Making Questionnaire (SDM-Q9). The secondary outcome was the change in anxiety level measured using the validated State-Trait Anxiety Inventory (STAI-6).

**Results:** A total of 51 patients, 28 in the 3D-printed arm and 23 in the standard care arm, met the inclusion criteria and agreed to participate. The mean age of participants was 51.1 years, with 55% female. Patients counseled with the 3D-printed model reported significantly higher involvement in shared decision making (mean score of 93.9 [SD 8.8] vs. 82.9 [SD 12.1], p <0.001), which was also clinically significant. Additionally, using a 3D-printed model significantly reduced anxiety scores (47.3 [6.0] to 46.4 [5.6]) compared to patients taught using conventional methods (48.3 [7.4] to 51.1 [9.7], p <0.008), but this effect was not deemed clinically significant.

**Conclusions:** The use of 3D models at pre-operative clinic visits improves shared decision making among patients planning to undergo colorectal surgery.

**Relevance:** This study highlights the potential of using 3D-printed models as an effective tool for a more collaborative and informed patient-clinician interaction. Given the positive findings, we recommend broadly implementing this technology.

**Trial registration:** clinicaltrials.gov Identifier: NCT06625008

**Key Points:** *Question:* Does pre-operative education aided by 3D-printed anatomic models improve the perceived shared decision making experience versus patients counseled using standard care practices?

*Findings:* This cluster randomized controlled trial compared the validated Shared Decision Making Questionnaire (SDM-Q9) scores among these study cohorts. The group counseled using 3D-printed models showed significantly higher levels of shared decision making than the group counseled using standard 2D images; this difference was also clinically meaningful.

*Meaning:* The use of 3D-printed models in preoperative counseling improves shared decision making when compared to the current practice of using 2D images.

## Introduction

Surgery is a life-altering event with permanent ramifications for the patients and their caregivers. Patients must receive satisfactory education regarding their disease process, treatment plans, and any alternatives to the proposed treatment. Poor pre-operative patient understanding can lead to lower compliance with pre-and post-operative care, as well as increased patient anxiety surrounding their disease and treatment ^1,2^. Shared decision making (SDM) involves patients and clinicians working together to select the most appropriate treatment option, considering patient preferences, values, and possible treatment outcomes ^3,4^. Patients involved in decision making are more satisfied and less anxious about their disease and potential treatment options ^5,6^.

Surgical patients report a poor understanding of their disease and treatment plan ^1,7,8^. Additionally, patients often report low involvement in their healthcare ^9^. Surgical professionals rank relaying information as the most essential factor for improving patient involvement^10^. While, physician-patient conversations and other conventional teaching methods, such as illustrations, can help inform patients. At times, it is insufficient, as it relies not only on a patient’s understanding of anatomy and physiology but also on imagining the disease and procedure superimposed on all this, all in a situation of duress ^1,7,11,12^. 3D printed models provide an adjunct to conventional consenting methods by improving patients’ understanding of their disease process and surgical options ^2,13–24^.

Little is known about the impact of 3D-printed models on shared decision making and patient anxiety levels in colorectal surgery. We hypothesized that patients considering colorectal surgery who are educated preoperatively using 3D-printed models will perceive increased levels of shared decision making and suffer lower procedural anxiety.

## Methods

### Design and Population

We conducted a single-center cluster randomized control trial in patients undergoing major abdominal colon or rectal surgery at an academic medical institution from March 2022 to June 2023. Patients were eligible if they were 18 years or older and were scheduled for surgical intervention that would involve partial or complete resection of the colon and/or rectum. Six consenting colorectal surgeons were randomly assigned to study arms using the opaque sealed envelope method ^25^. All consenting patients were assigned based on physician availability.

Participant enrollment, assignment to study arms, and cluster randomization were performed by persons unrelated to the research. The cluster randomization method was used to prevent the mixing and contamination of the intervention. Blinding was not performed as it was not feasible. This study protocol was approved by the institutional review board (IRB# 211314), and it was conducted per the current version of the Declaration of Helsinki ^26^ and in agreement with the International Conference on Harmonization guidelines on Good Clinical Practice ^27^. Patients provided informed consent at the time of enrollment. This trial was registered at ClinicalTrials.gov (NCT06625008). The current trial followed the Consolidated Standards of Reporting Trials 2010 (CONSORT) guidelines.

### Sample Selection/Power Calculation

As there is no established data regarding SDM-Q-9 efficacy in decision making in colorectal surgery, we used the mean and standard deviation of SDM-Q-9 available in the literature [86.7 (±11.6)]^28^ to calculate our sample size. To ensure that this study had adequate power to detect a clinically meaningful effect size and provide data for future studies, we assumed a 5% risk of type I error and a 20% risk of type II error, a 1:1 enrollment ratio, and a targeted increase of 3 times the minimal clinically important difference (4 ^29^). This meant that 20 patients needed to be allocated to each arm. Since this intervention has not been utilized in colorectal patients before, we targeted an expected rejection rate of 50%, meaning 80 patients were approached (40 in each arm).

### Study arms

Six surgeons and 51 patients participated in this study. Each surgeon was randomized to conduct relevant pre-operative patient teaching and to consent for upcoming surgery using either the “3D-printed model” or the “standard care” (usual care, consisting of 2D images and verbal explanations of disease and treatment options) (Figure 1). Surgeons in the intervention arm were briefed about the 3D-printed model and its development to allow them to use it seamlessly in clinical interactions. However, they were not informed of the perceived benefits to prevent introducing a bias.

**Figure 1.**
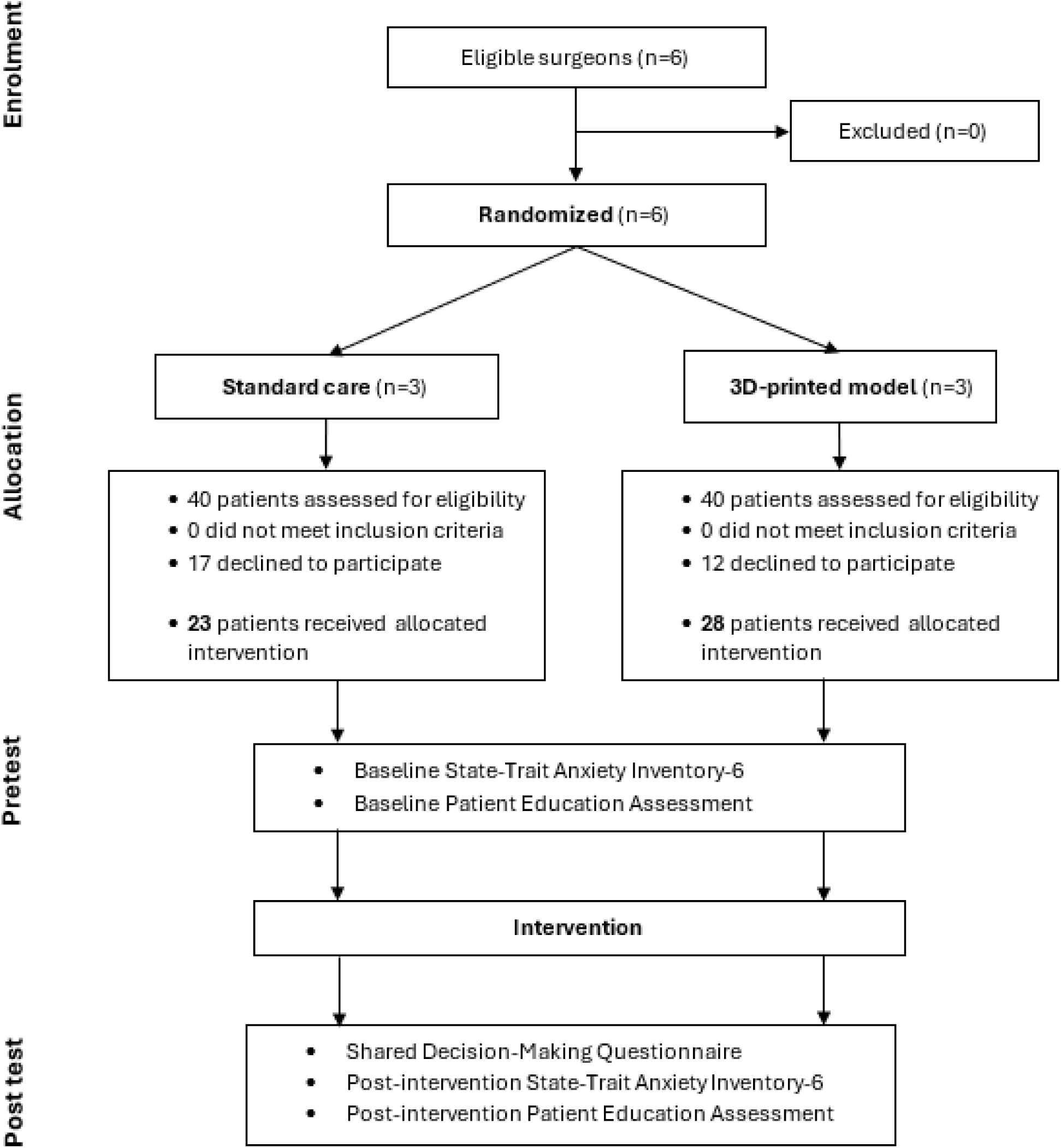
CONSORT flow diagram.

### Development of the 3D model

In collaboration with the Department of Radiology, the authors designed and developed a 3D-printed human torso model with a colon and rectum. The modular design allowed each segment of the colon and rectum to be magnetically detached and reattached, allowing effective teaching of patients on colonic segments undergoing surgery. The model included a representation of a stoma on the abdominal wall, allowing patients to appreciate its appearance and function (Figures 2 A-D). The code to print this 3D model can be requested from the corresponding author on reasonable request.

**Figure 2A.**
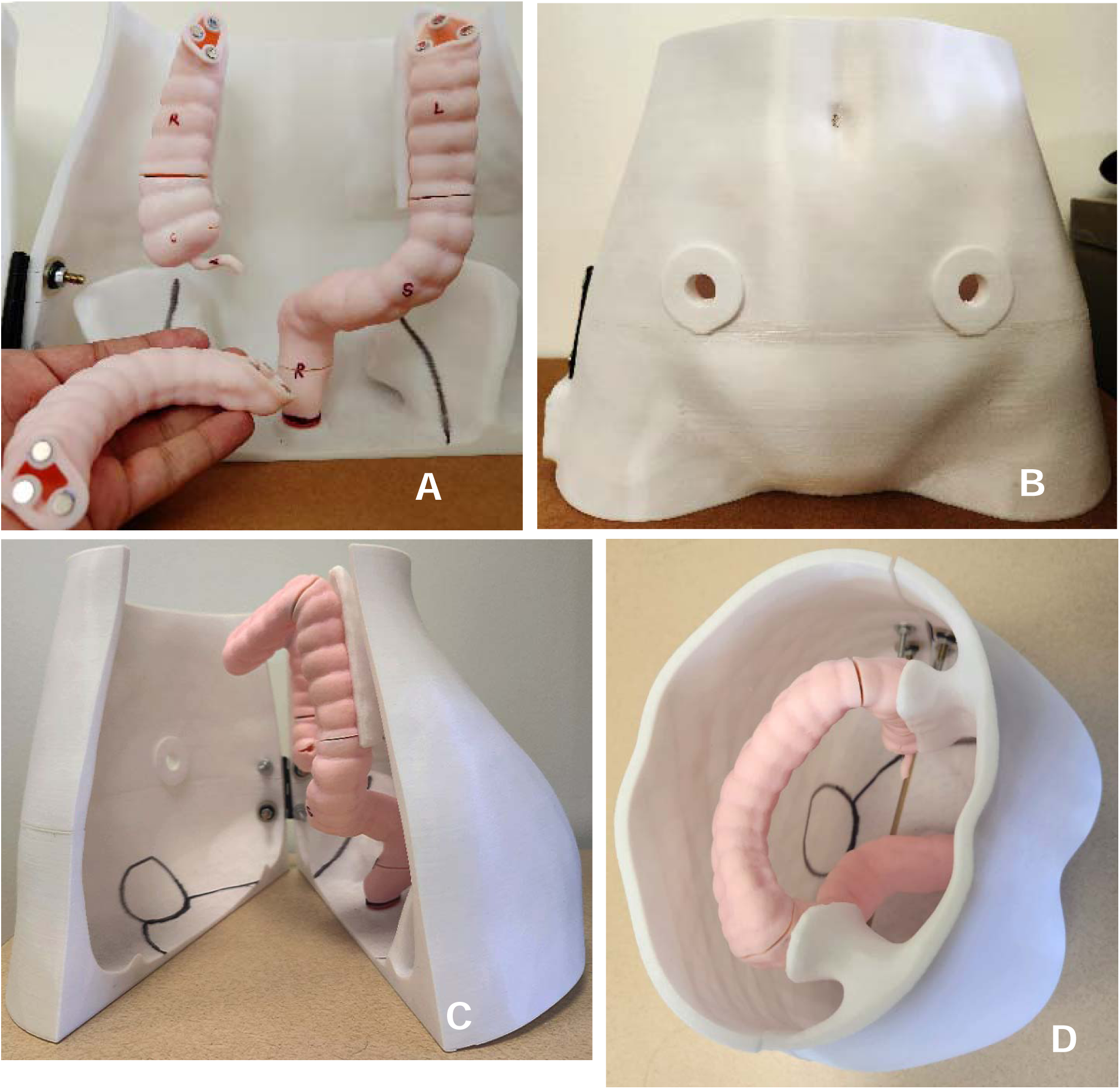
3D-printed model with detachable magnetic modules. Figure 2B. 3D-printed model showing ostomy sites. Figure 2C. 3D-printed model showing the relation of the colon and rectum to the urinary system in an open model. Figure 2D. A top-down view of the 3D-printed model showing the relation of the colon and rectum to the urinary system in a closed mode.

### Primary and Secondary Study Outcome Measures

Consenting participants who fulfilled the inclusion criteria were assessed for the knowledge of their disease and anxiety levels before and after the intervention (pre-operative consent and education). Additionally, patient health literacy using the 4-question validated BRIEF Health Literacy screening tool ^30,31^ was measured prior to the intervention, and their perceived involvement in the decision making process was measured once post-intervention.

The validated Shared Decision Making Questionnaire (SDM-Q-9) ^32^ was employed to assess patient-perceived involvement in decision making alongside their surgeon. Based on previous literature, a minimal clinically important difference (MCID) for SDM-Q-9 is 4 points ^29^. Patient health literacy (in English) was measured using the validated and self-administered BRIEF: Health Literacy Screening Tool ^30^, a 4-question survey that helps gain insight into a patient’s ability to understand medical interactions and information. As per the tool guide, health literacy scores were characterized as inadequate (<12), marginal (13-16), and adequate (17-20). Patient anxiety and stress levels were measured using the self-administered, validated six-item State-Trait Anxiety Inventory (STAI-6) ^33^, and a ten-unit change was considered the MCID^34^. The change in patient knowledge before and after the intervention was measured by the Patient Education Assessment questionnaire, developed by the study authors after literature review, expert consultation, and beta testing among a similar patient cohort to assess understanding and respondent burden. This education questionnaire comprised eight items divided evenly between understanding the disease and the surgery and complication risks (Annexure 2).

### Statistical Analysis

Continuous variables were reported as mean [standard deviation (SD)], while frequency (%) was reported for categorical variables. Patient characteristics were compared between the 3D-printed model and standard care arms with the chi-squared test for categorical variables and the Student’s t-test for comparisons between continuous variables. SDM-Q-9, STAI-6, and Patient Education Assessment mean scores were compared between the 3D-printed model and standard care arms using the Student’s t-test. Post-intervention STAI-6 and Patient Education Assessment mean scores were also compared using the Analysis of Covariance (ANCOVA) test. The post-intervention score was treated as a dependent variable, while the pre-intervention score was treated as a covariate. ANCOVA with pre-intervention scores as a covariate provides a more appropriate and informative analysis when compared with repeated measures of ANOVA ^35^. A similar subgroup analysis was conducted after dichotomizing health literacy into adequate (17-20) and inadequate/marginal (<17) groups. The level of significance was set at α = 0.05 throughout the manuscript. Statistical analyses were performed using Stata (version 17, StataCorp LLC, College Station, TX).

## Results

### Comparison of baseline characteristics

Fifty-one patients undergoing surgery by six different surgeons were prospectively enrolled in this study. After cluster randomization, 28 participants were enrolled in the intervention arm (using a 3D model for preoperative counseling), while 23 participants were enrolled in the control arm utilizing the standard care of using 2D images. Baseline patient demographic and health literacy data stratified by study arms can be seen in Table 1. The mean age of study participants was 51.1 years, with no significant differences between the 3D and standard care arms. Fifty-five percent of patients were female, and most self-identified as White (86%), with 8% identifying as Black and 2% as Other. The highest degree of education for most patients was a high school diploma/GED (44%), followed by a graduate degree of more significant (26%) and an undergraduate degree (24%). Six percent of patients reported less than a high school diploma. Using the self-reported BRIEF: Health Literacy Screening Tool, 15.7% percent of the patients had inadequate health literacy, while 21.6 % had marginal health literacy; adequate health literacy was seen amongst 62.7%. There was no significant distribution difference in healthy literacy across study arms (Table 1). Furthermore, no significant differences in baseline STAI-6 scores and Patient Education Assessment were noted (Table 2).

**Table 1:** Patient characteristics, stratified by Standard care and 3D-printed model arms.

| Participant characteristics | Standard care (n=23) | 3D model (n=28) | p-value |
| --- | --- | --- | --- |
| Age, mean ± SD, years | 55.3 (14.0) | 47.7 (14.3) | 0.06 |
| Sex, n (%) |  |  | 0.44 |
| Female | 14 (61%) | 14 (50%) |  |
| Male | 9 (39%) | 14 (50%) |  |
| Race, n (%) |  |  | 0.60 |
| White | 20 (87%) | 24 (86%) |  |
| Black | 1 (4%) | 3 (11%) |  |
| Other | 1 (4%) | 0 (0%) |  |
| Prefer not to answer | 1 (4%) | 1 (4%) |  |
| Ethnicity, n (%) |  |  | 0.65 |
| Hispanic | 0 (0%) | 1 (4%) |  |
| Non-Hispanic | 22 (96%) | 26 (93%) |  |
| Unknown | 1 (4%) | 1 (4%) |  |
| Education, n (%) |  |  | 0.75 |
| Less than a high school diploma | 1 (5%) | 2 (7%) |  |
| High school diploma/GED | 10 (45%) | 12 (43%) |  |
| Undergraduate degree | 4 (18%) | 8 (29%) |  |
| Graduate degree or greater | 7 (32%) | 6 (21%) |  |
| BRIEF Health Literacy Score, n (%) |  |  | 0.69 |
| <12 | 4 (17%) | 4 (14%) |  |
| 13–16 | 6 (26%) | 5 (18%) |  |
| 17–20 | 13 (57%) | 19 (68%) |  |

Note:
SD standard deviation

**Table 2:** Mean Scores for Patient Education Assessment, STAI-6, and SDM-Q9 for the Standard care versus 3D-printed model arms, overall and stratified by BRIEF Health Literacy score.

|  | Categories | Standard care | 3D model | p-value |
| --- | --- | --- | --- | --- |
| Overall | Post-Intervention SDM-Q9 | 82.9 (12.1) | 93.9 (8.8) | **<0.001**<sup>a</sup> |
|  | Baseline STAI-6 | 48.3 (7.4) | 47.3 (6.0) | 0.597<sup>a</sup> |
|  | Post-Intervention STAI-6 | 51.1 (9.7) | 46.4 (5.6) | **0.008**<sup>b</sup> |
|  | Baseline Patient Education Assessment | 56.8 (26.7) | 63.8 (21.3) | 0.306<sup>a</sup> |
|  | Post-Intervention Patient Education Assessment | 86.9 (13.7) | 83.9 (15.2) | 0.318<sup>b</sup> |
| BRIEF < 17 | Baseline Patient Education Assessment | 60.0 (29.9) | 56.9 (15.5) | 0.787<sup>a</sup> |
|  | Post-Intervention Patient Education Assessment | 86.3 (12.4) | 84.7 (18.5) | 0.930<sup>b</sup> |
|  | Baseline STAI-6 | 51.0 (8.3) | 45.6 (6.7) | 0.137<sup>a</sup> |
|  | Post-Intervention STAI-6 | 52.6 (9.1) | 45.2 (5.6) | 0.133<sup>b</sup> |
|  | Post-Intervention SDM-Q9 | 80.0 (13.3) | 93.3 (7.7) | **0.017**<sup>a</sup> |
| BRIEF ≥ 17 | Baseline Patient Education Assessment | 54.2 (24.6) | 67.1 (23.3) | 0.151<sup>a</sup> |
|  | Post-Intervention Patient Education Assessment | 87.5 (15.6) | 83.6 (13.9) | 0.308<sup>b</sup> |
|  | Baseline STAI-6 | 46.2 (6.1) | 48.1 (5.7) | 0.370<sup>a</sup> |
|  | Post-Intervention STAI-6 | 50.0 (10.3) | 47.0 (5.7) | **0.038**<sup>b</sup> |
|  | Post-Intervention SDM-Q9 | 85.0 (11.3) | 94.1 (9.4) | **0.011**<sup>a</sup> |

Note:
<sup>a</sup> Compared using the Student's t-test<sup>b</sup> Compared using the Analysis of Covariance (ANCOVA) test, after adjusting for baseline scores
Quantitative variables are reported as mean (SD)
Bold p-values are significant at p<0.05
STAI-6 Spielberger State-Trait Anxiety Inventory, SDM-Q9 Shared Decision-Making Questionnaire, SD standard deviation

### Primary Outcome

The SDM-Q-9 survey scores for the 3D-printed model arm were significantly higher than the standard care arm (93.9 vs. 82.9, p <0.001), more than twice the established four-unit MCID, demonstrating a clinically significant increased shared decision making amongst patients and providers (Table 2) (Figure 3A). Subgroup analysis based on dichotomizing health literacy (BRIEF Health Literacy Score <17 vs. ≥17) yielded a similar trend in SDM-Q-9 scores with health literacy scores <17 (93.3 vs. 80.0, p=0.017) as well as ≥17 (94.1 vs. 85.0, p=0.011) (Table 2).

**Figure 3A.**
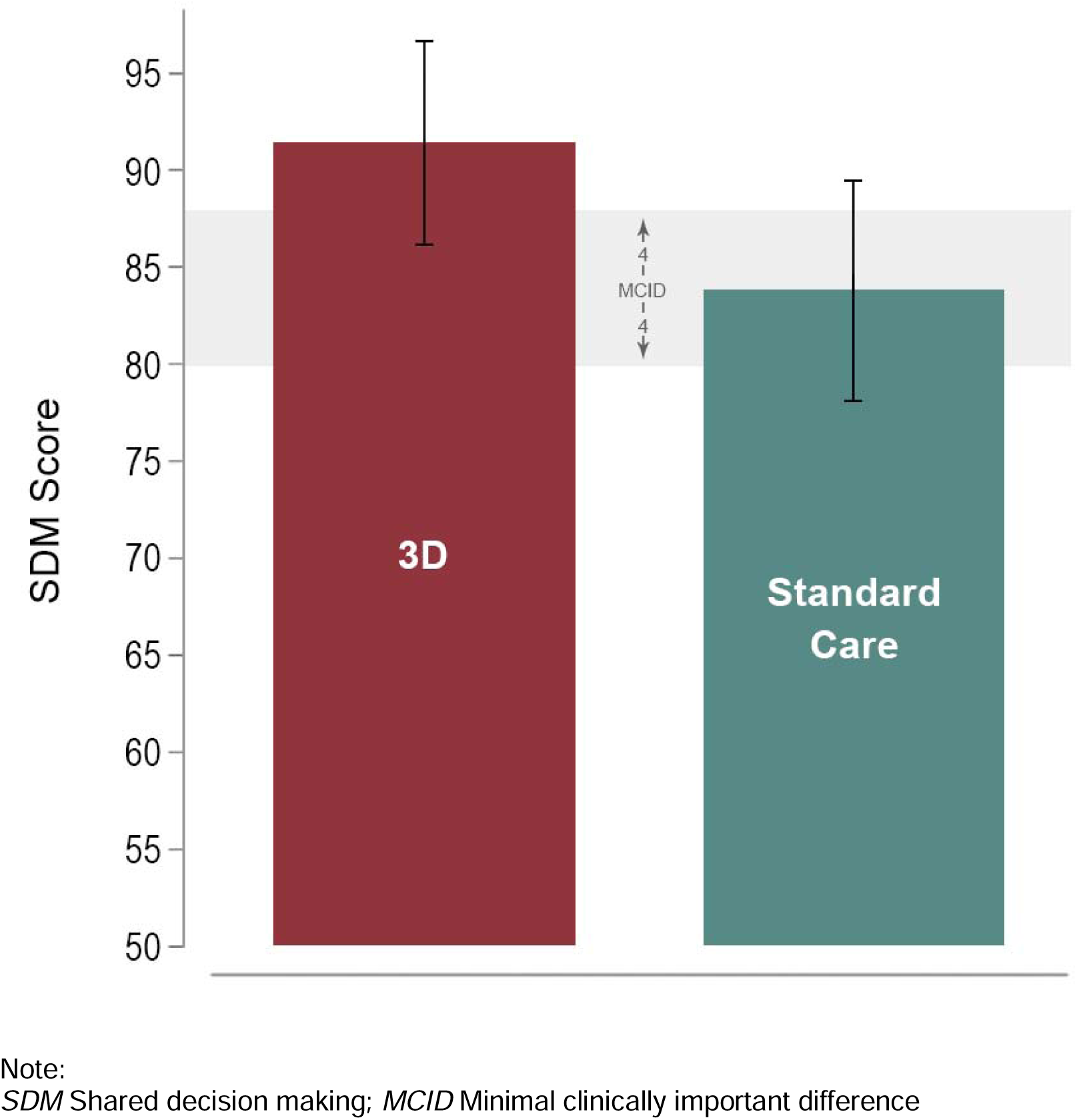
Post-visit shared decision-making scores using the SDM-Q-9 questionnaire, compared between the 3D-printed model and standard care arm.

### Secondary Outcomes

Post-intervention anxiety scores increased in the standard care arm (48.3 to 51.1) but decreased (47.3 to 46.4) in the 3D-printed model arm (Table 2) (Figure 3B); a similar trend was seen when dichotomized by health literacy. After adjusting for baseline scores, patients within the 3D-printed model arm had a significant reduction in anxiety levels of 0.9 points, compared to the standard care arm, in which the anxiety increased by 2.8 points (p = 0.008) (Table 2). After dichotomizing by health literacy, a significant reduction of 1.1 points was only seen with a health literacy of ≥17 in STAI-6 scores among patients in the 3D-printed model arm vs. standard care arm, where anxiety increased by 3.8 points (p=0.038). However, the MCID threshold of ten points was not crossed, meaning a clinically significant difference was not seen (Table 2).

**Figure 3B.**
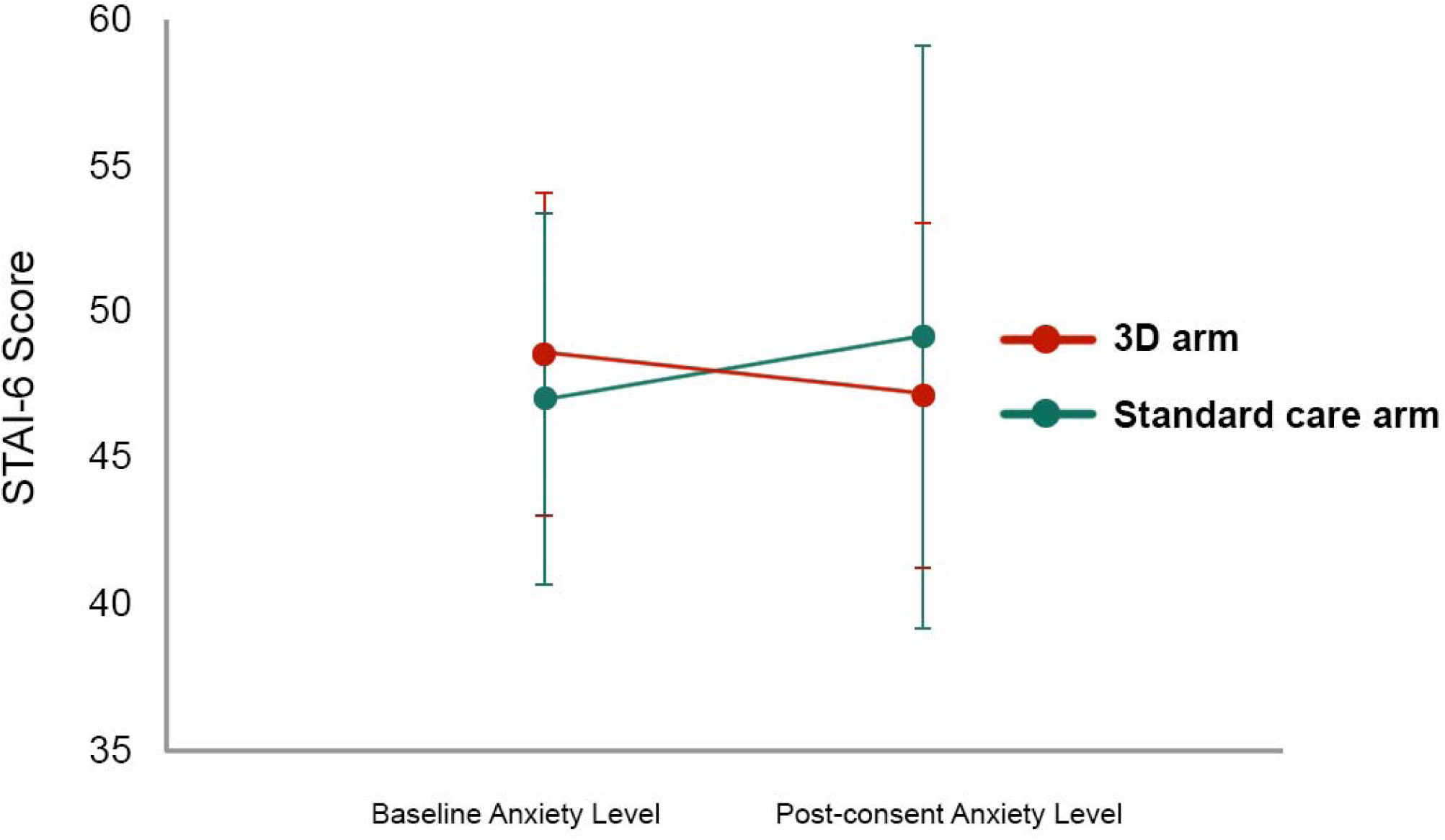
Impact of 3D-printed model arm on patient anxiety.

While both arms had an improvement in the overall Patient Education Assessment score (30.1% for the standard care arm and 20.1% in the 3D-printed model arm), there was no significant difference between the two arms after adjusting for baseline scores regardless of patient health literacy (i.e., adequate vs. inadequate/marginal) level (Table 2). Across the study arms, there was a significant improvement in understanding the locations of disease and resection sites. Patients in the 3D-printed model arm had a significant improvement in the understanding of potential anatomic structures at risk of damage during the operation (p=0.016). At the same time, the standard care helped improve an understanding of the type of incisions among the patients (p=0.008) (Supplemental Table 1).

## Discussion

Our study is the first randomized trial to compare the effectiveness of a 3D-printed model versus standard of care on quality of decision making, patient-provider engagement, anxiety, and patient education in patients undergoing colorectal surgery. The use of a 3D-printed model improved the decision making process regardless of patient health literacy. Patients who were educated using 3D-printed models reported decreased anxiety after their surgical consultation compared to patients taught using standard care. These findings demonstrate the effectiveness of this 3D-printed model in aiding patient education during the preoperative consent before colorectal surgery.

The pre-operative clinic visit is a complex and anxiety-provoking experience for patients^36^. A thorough understanding of their condition, management options, and prognosis in the pre-operative setting is critical. From an ethical standpoint, patients must have a sufficient understanding of their condition, the surgical plan, and expected post-operative recovery to provide informed consent. In turn, optimal comprehension may result in increased ‘buy-in’ or a sense of shared decision making from patients and, ultimately, more effective healthcare delivery and adherence to the recommended care plan ^37–40^. Poor patient comprehension continues to pose significant challenges in surgery, and optimal patient education strategies may improve patient anxiety regarding surgery. In this regard, devising methods to improve patient education in the pre-operative setting is critical. With the rapid rise of 3D-printing technology in the 21st century, using models in this context may be a potential solution ^14,19–24,41–45^. Several studies have also created patient-specific 3D models as part of the patient education process; however, resource and time intensiveness have reduced its practicality for routine use ^18,46^. This concept has recently been extended to surgical training, with studies noting improved outcomes ^37–42,47^.

A patient’s sense of shared decision making is critical in developing a positive patient-physician relationship and may improve patient adherence to care plans and ultimately improved long-term outcomes. Shared decision making in pre-operative care is a process during which surgeons and patients cooperate to form care plans that comply with patients’ goals and values, thus ensuring patient autonomy and patient-centered care ^48^. In the present study, patients had a significantly greater sense of shared decision making in the 3D-printed model arm, regardless of patient health literacy. These results indicate that patients educated using a 3D-printed model arm felt more involved in the consultation process and had a greater say in their care plan. This can be extrapolated to improved patient satisfaction, as Zheng et al. demonstrated ^16^. We believe utilizing a shared, tangible reference was the key factor in improving communication between the patient and provider, allowing for a significant improvement in shared decision making.

Our study was effective in showing that 3D-printed model-assisted teaching in the pre-operative consent process can help reduce anxiety ahead of surgery. Using the STAI-6 survey, Biro et al. also found similar results prior to Mohs surgery with the implementation of 3D-printed models ^49^. Albeit using different scales, Demirel et al., in their cross-sectional study, demonstrated that lower pre-operative anxiety was observed with a higher health literacy score ^50^. Literature iterates that pre-operative anxiety is associated with increased post-operative morbidity ^51–53^. These findings emphasize the need for strategies that effectively reduce patient anxiety in the pre-operative setting to improve post-operative outcomes.

In the present study, the post-intervention Patient Education Assessment scores improved with patient education and consent; however, no statistically significant difference was observed between the 3D-printed model and standard care arms. In fact, among the Patient Education Assessment items, only the items pertaining to the location of the disease and resection demonstrated significant improvement in the proportion of correct answers in both arms. Conversely, only the 3D-printed model arm improved understanding of the anatomic structures at risk of damage during surgery. This may be due to improved visualization of the overall anatomy (of the region) and the proximity of adjacent structures. Similarly, Wake et al. found that 3D models helped learn about the anatomy, disease, cancer location, and treatment plan ^23^. Patients in the standard care arm significantly improved their understanding of the incision type compared to the 3D-printed model arm. This is probably because it is easier to demonstrate incisions on 2D images than a standard 3D model. These results demonstrate improved patient understanding regardless of education technique, which persisted regardless of baseline patient health literacy. Currently, the literature is conflicting on this front, with Crepeau et al. demonstrating unexpectedly poor understanding and recall for elective orthopedic surgery ^7^ while Kim et al. demonstrated that using patient-specific 3D-printed models improved pre-operative patient education ^19^. These questionnaires are often not validated or do not cover a broad range of procedures. This highlights the need to develop and validate patient education questionnaires and 3D models that are specialty-specific, disease-specific, or patient-specific and their subsequent use in future randomized control trials to understand the full effect of 3D-printed models in patient education.

### Limitations

Since this study is conducted at a single large urban tertiary care hospital, the results might not apply to disparate practices. Second, the results of this study are only applicable to patients fluent in the English language owing to the language and validation of the scales used. Another limitation of the study was the lack of ethnic diversity among the patients. We plan to address this in the future multi-institutional trial to study the effects of such patient-reported outcome measures across a spectrum of diseases and patients. A possible response shift bias was not evaluated; however, ANCOVA analysis could partly reduce the effect. The current study did not account for time differences in counseling for each arm, which can be a decisive factor in adopting 3D-printed models in clinical practice.

### Future considerations

Including patient satisfaction, post-operative outcomes, knowledge retention, qualitative analysis of shared decision making, and surgeons’ perspective of shared decision making can help better understand the effectiveness of the intervention and patient perspective. Furthermore, disease-specific models might help enhance patient understanding compared to general models. In this study, the knowledge of the type of incision did not significantly improve in the 3D-printed model arm compared to the standard care arm, this can be a point of improvement in the design of 3D-printed models.

### Conclusion

The use of 3D-printed models for consent of elective surgeries improved shared decision making and reduced anxiety among patients undergoing elective colorectal surgeries. However, overall patient education remained similar to standard practice. Using a 3D-printed model for pre-operative patient counseling had benefits over standard care and was not associated with the worsening of any patient-reported outcome measure; as such, we recommend using it.

## Data sharing statement

Individual participant data will not be made available, but researchers who provide a methodologically sound proposal approved by an independent review committee can direct their requests to the corresponding author.

## Acknowledgments

None

