## Supplementary Table 1 for "Impact of 3D Printed Models on Shared Decision Making: A Cluster Randomized Controlled Trial"

**Supplemental table 1:** Percentage of correct responses for each Patient Education Assessment Item for the Standard care versus 3D-printed model arm.

|  | **% Correct** | | **Δ % Correct** | **p-value** ^a^ |
| --- | --- | --- | --- | --- |
| **Categories** | **Baseline** | **Post-Intervention** |  |  |
| **Standard care (n=23)** |  |  |  |  |
| Is the colon a hollow organ? | 82.6 | 95.2 | 12.6 | 0.50 |
| Which part of your colon contains the disease? | 22.7 | 76.2 | 53.5 | **0.001** |
| Which part of your colon will the surgeon remove? | 27.3 | 71.4 | 44.2 | **0.004** |
| What type of incision will be used? | 47.8 | 86.4 | 38.5 | **0.008** |
| Will you have an ostomy (bag) after the surgery? | 69.6 | 95.5 | 25.9 | 0.063 |
| What is an anastomotic leak? | 73.9 | 90.9 | 17.0 | 0.25 |
| What is a potential complication of your surgery? | 87.0 | 95.5 | 8.5 | 1.00 |
| What can be damaged during surgery? | 52.2 | 72.7 | 20.6 | 0.34 |
| **3D-printed model (n=28)** |  |  |  |  |
| Is the colon a hollow organ? | 89.3 | 96.4 | 7.1 | 0.50 |
| Which part of your colon contains the disease? | 32.1 | 71.4 | 39.3 | **0.001** |
| Which part of your colon will the surgeon remove? | 25.0 | 75.0 | 50.0 | **<0.001** |
| What type of incision will be used? | 71.4 | 78.6 | 7.1 | 0.75 |
| Will you have an ostomy (bag) after the surgery? |  | 92.9 | 10.7 | 0.38 |
| What is an anastomotic leak? | 78.6 | 92.9 | 14.3 | 0.22 |
| What is a potential complication of your surgery? | 82.1 | 89.3 | 7.2 | 0.63 |
| What can be damaged during surgery? | 50.0 | 75.0 | 25.0 | **0.016** |
| Note:  ^a^ Comparison between the percentage of correct answers before and after counseling with standard care arm or 3D-printed model arm  Bold p-values are significant at p<0.05 | | | | |
